# Single-cell RNA Analysis on ACE2 Expression Provides Insight into SARS-CoV-2 Blood Entry and Heart Injury

**DOI:** 10.1101/2020.03.31.20047621

**Authors:** Jieyu Guo, Xiangxiang Wei, Qinhan Li, Liliang Li, Zhaohua Yang, Yu Shi, Yue Qin, Xinyue Zhang, Xinhong Wang, Xiuling Zhi, Dan Meng

**Author notes:** Corresponding authors. (D. M.). Jieyu Guo, Xiangxiang Wei, and Qinhan Li contributed equally to this work.

## Abstract

COVID-19 is a global pandemic with high infectivity and pathogenicity, accounting for tens of thousands of deaths worldwide. Recent studies have found that the pathogen of COVID-19, SARS-CoV-2, shares the same cell receptor Angiotensin converting enzyme II (ACE2) with SARS-CoV. The pathological investigation of COVID-19 death showed that the lung had the characteristics of pulmonary fibrosis. However, how SARS-CoV-2 spreads from the lungs to other organs has not yet been determined. Here, we performed an unbiased evaluation of cell-type specific expression of ACE2 in healthy and fibrotic lungs, as well as in normal and failed adult human hearts, using published single-cell RNA-seq data. We found that ACE2 expression in fibrotic lungs mainly locates in arterial vascular cells, which might provide the route for bloodstream spreading of SARS-CoV-2. The failed human hearts have a higher percentage of ACE2-expressing cardiomyocytes, and SARS-CoV-2 might attack cardiomyocytes through the bloodstream in patients with heart failure.

Moreover, ACE2 was highly expressed in cells infected by RSV or MERS-CoV and in mice treated by LPS. Our findings indicate that patients with pulmonary fibrosis, heart failure, and virus infection have a higher risk and are more susceptible to SARS-CoV-2 infection. SARS-CoV-2 might attack other organs by getting into the bloodstream. This work provides new insights into SARS-CoV-2 blood entry and heart injury and might propose a therapeutic strategy to prevent patients from developing severe complications.

## Introduction

COVID-19 is a global pandemic with high infectivity and pathogenicity, accounting for tens of thousands of deaths worldwide. The pathogen named severe acute respiratory syndrome coronavirus 2 (SARS-CoV-2) showed 79.5% genome similarity to SARS-CoV and 96% to a bat coronavirus RaTG13^1^. Angiotensin converting enzyme II (ACE2) can specifically bind to the S1 domain of SARS-CoV S protein as a functional receptor^2^ and is also the cellular receptor of SARS-CoV-2. ACE2 has been widely recognized as a negative regulatory enzyme of the renin-angiotensin system (RAS) that converts Ang II into Ang1-7 /Ang 1-9 and has a protective role in cardiovascular function^3^. Upon spike protein of coronavirus binding to the cellular receptor ACE2, the type II transmembrane serine protease (TTSP) TMPRSS2 cleaves the SARS-CoV-2 spike protein to promote fusion of viral and cellular membranes^4^. FURIN also cleaves SARS-CoV-2 spike protein^5^. Both TMPRSS2 and FURIN play an essential role in viral entry^4,5^.

ACE2 was shown to be present in human lung alveolar epithelial cells^6^. Analysis of single-cell RNA-Seq (scRNA-seq) data in normal human lung tissues demonstrated that ACE2 is mainly expressed in type II alveolar cells (AT2)^7^. The pathological investigation of COVID-19 fatal cases showed exudation and hemorrhage, epithelium injuries, infiltration of macrophages, and fibrosis in the lungs^8^. Pulmonary fibrosis is not only due to that SARS-CoV-2 directly attacking the lung as its target organ, while the activation of cytokine storm in COVID-19 patients also contributes^9,10^. A retrospective cohort study indicates that patients with chronic obstructive pulmonary disease (COPD) and bilateral pulmonary infiltration are associated with higher mortality^11^. It may be due to the change of *ACE2* gene expression profile in lung tissues in patients with lung diseases, which makes these patients more susceptible to SARS-CoV-2 infection, but the cell-type specific expression change of ACE2 in healthy populations and patients with underlying lung diseases has not been reported.

The epidemiology analysis of COVID-19 has identified cardiac injury as one of the most severe organ function damages, with symptoms of arrhythmia and cardiac arrest^9^. Elevated high sensitivity Troponin I (hs-cTnI) or new ECG / echocardiographic abnormalities suggested that cardiac injury was present in 7.2% of patients and 22% of those who required ICU care^12^. Moreover, advanced age and preexisting cardiovascular diseases were high-risk factors of infection and lethality^13^. In a study involving 138 COVID-19 patients, many had cardiovascular-related comorbidities and/or complications, including severe hypertension (31.2%), cardiovascular disease (14.5%), and arrhythmia (16.7%)^12^. However, how SARS-CoV-2 spreads from the lungs to other organs and the cause of the high mortality rate in COVID-19 patients with preexisting cardiovascular diseases are still not fully understood.

To investigate the possible route of SARS-CoV-2 spreading from the lungs to other organs, we performed an unbiased evaluation of cell-type specific expression of ACE2 in healthy and fibrotic lungs, as well as in normal and failed adult human hearts, using published single-cell RNA-seq data. We found that ACE2 is mainly expressed in arterial vascular cells in fibrotic lungs, which might be essential for SARS-CoV-2 to enter the bloodstream and to start its circulatory spreading. We observed higher enrichment of ACE2 expression in cardiomyocytes of failed adult human hearts, and higher gene expression of *ACE2* in cells infected by the respiratory syncytial virus (RSV) or middle east respiratory syndrome-coronavirus (MERS-CoV). Our findings indicate that patients with pulmonary fibrosis, heart failure, and virus infection have a higher risk and are more susceptible to SARS-CoV-2 infection.

## Results

### *ACE2* is mainly expressed in arterial vascular cells in fibrotic lungs

To explore whether there is a cell-type specific expression difference of the *ACE2* gene in healthy populations and patients with pulmonary fibrosis, we analyzed a published dataset containing scRNA-seq data of the normal whole-lung tissues from three healthy donors and fibrotic lung samples from three patients (GSE132771). 11594 cells in normal lung tissues and 13069 cells in fibrotic lung tissues were analyzed. We classified the entire population of normal lung tissue into five major cell types, including type I and type □ alveolar cells (AT1, AT2), ciliated bronchial epithelial cells, immune cells, fibroblasts and vascular cells based on their respective molecular features (Fig. S1A-S1B). Similar cell types were also observed in fibrotic lung samples (Fig. S1C-S1D). We found that the majority of ACE2-expressing cells in normal human lung tissues were AT2 cells (*SFTPC*-positive) (Fig. 1A and 1C), which is consistent with a previous study^7^. Unexpectedly, ACE2 was mainly expressed in arterial vascular cells, but rarely in AT2 in fibrotic lung tissues (Fig. 1B and 1D), indicating that the distribution of ACE2 in fibrotic lung tissues has shifted compared to the normal.

**Figure 1.**
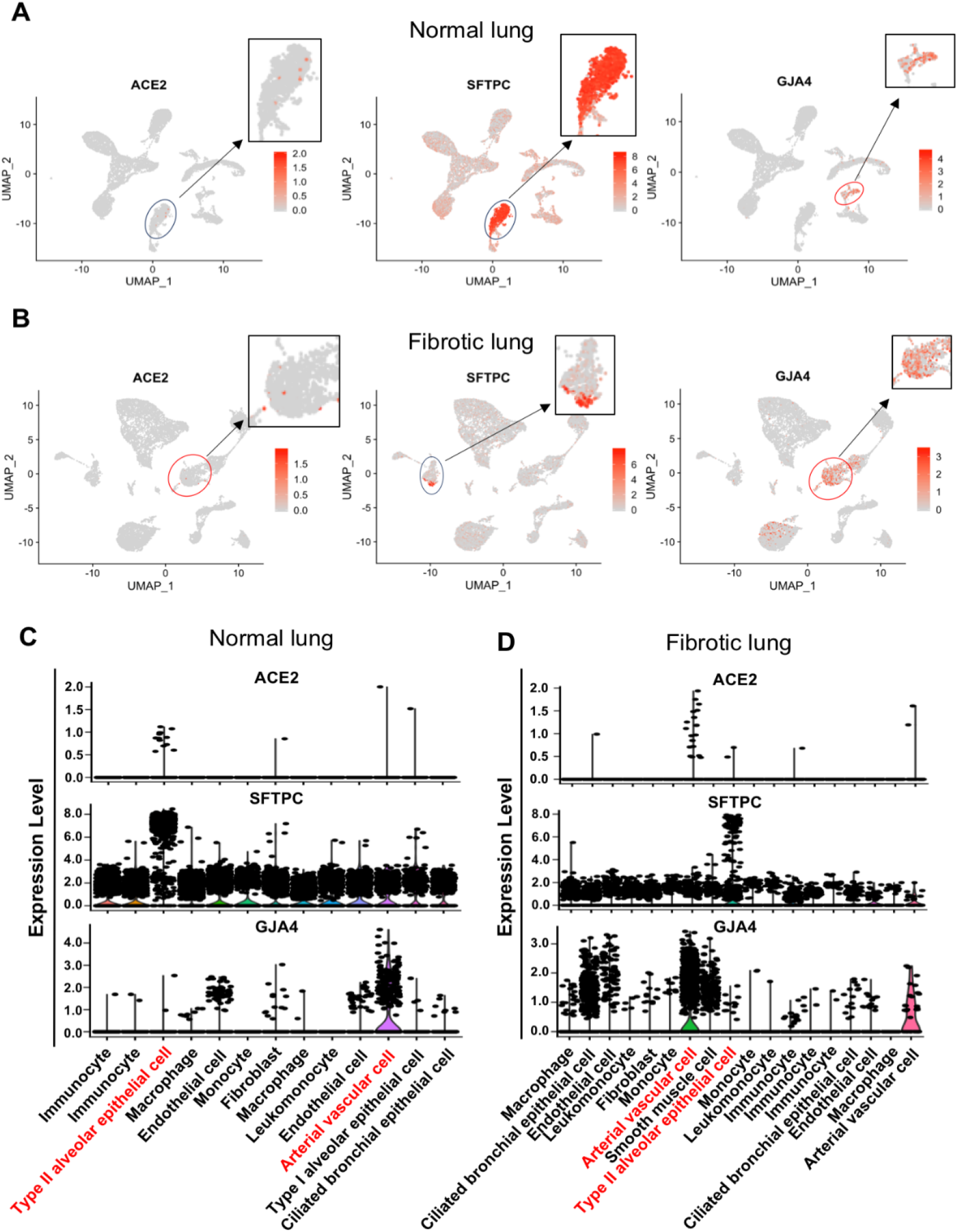
Different distribution of ACE2-expressing cells between normal and fibrotic lungs. A and B, Feature plot showing the distribution of *ACE2, SFTPC* and *GJA4* expression levels in normal lungs (A) and in fibrotic lungs (B). C and D, Violin plot showing the distribution of *ACE2, SFTPC* and *GJA4* expression levels in normal lungs (C) and in fibrotic lungs (D).

Further analysis showed that chemokines (*CCL2, CXCL12*) and complement component (*C1R*) were highly expressed in *ACE2*-positive arterial vascular cells of fibrotic lungs, but showed lower expression in *ACE2*-positive AT2 cells of normal lungs (Fig. 2A-2B), which suggests that *ACE2*-expressing arterial vascular cells in fibrotic lungs may be associated with inflammation. SARS-CoV-2 depends on TMPRSS2 and FURIN to infect target cells ^4,5^. Both TMPRSS2 and FURIN were expressed in AT2 cells of normal lungs, whereas only FURIN was expressed in arterial vascular cells of fibrotic lungs (Fig. 2A-2B), indicating that the entryway of SARS-CoV-2 might depend on FURIN in fibrotic lungs, which is different from the normal lungs. These observations show that *ACE2* is mainly expressed in arterial vascular cells in fibrotic lungs, which might help SARS-CoV-2 to enter blood vessels and cause bloodstream spreading of SARS-CoV-2 after infection.

**Figure 2.**
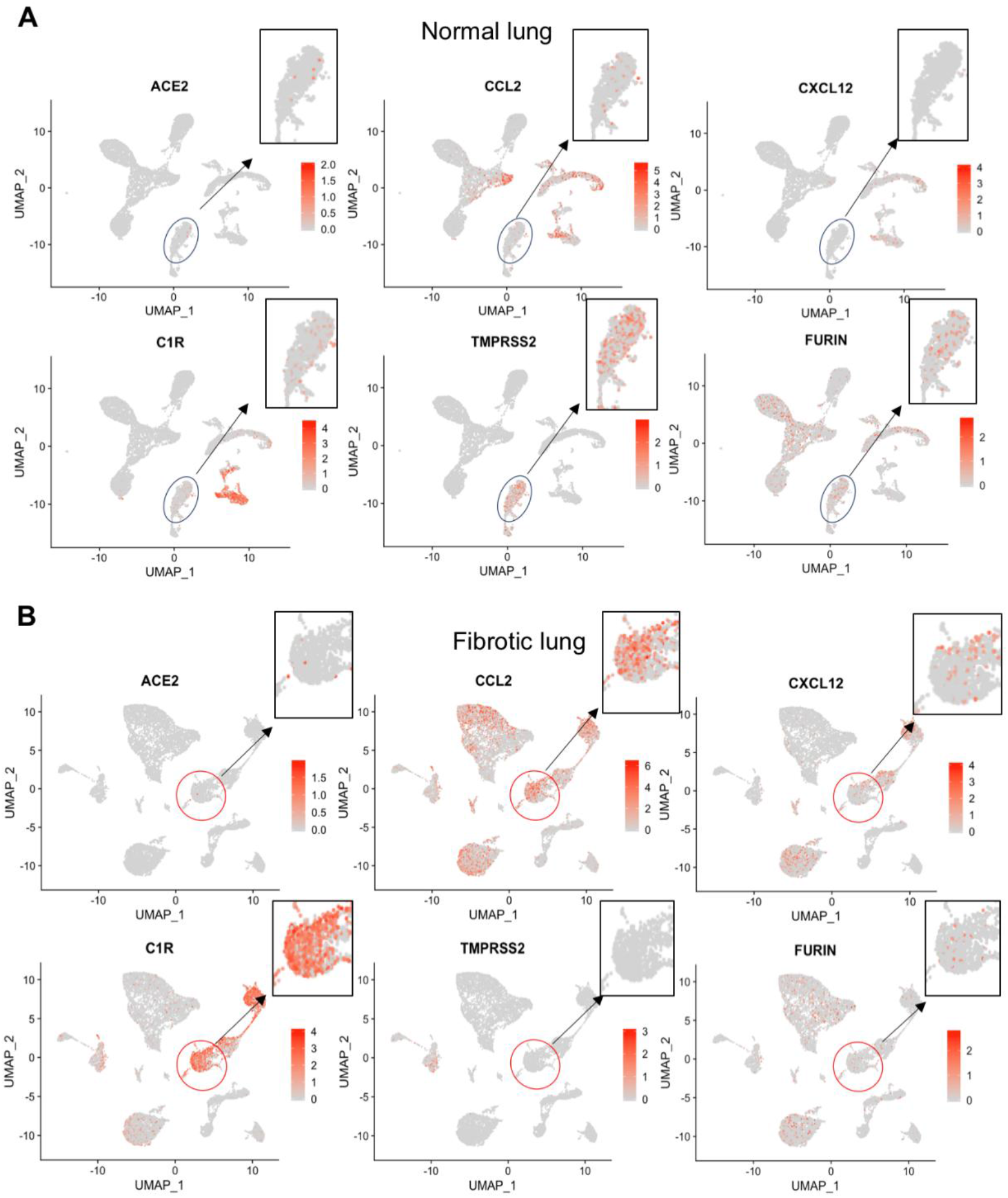
Distribution of chemokine and proteases between normal and fibrotic lungs. A and B, Feature plot showing the distribution of *ACE2, CCL2, CXCL12, C1R, TMPRSS2* and *FURIN* expression levels in normal lungs (A) and in fibrotic lungs (B).

### Failed human hearts have a higher percentage of ACE2-expressing cardiomyocytes

To assess the cell-type specific expression of *ACE2* gene in heart, we analyzed the published scRNA-seq data of adult human heart tissues from 12 healthy donors (GSE109816) and six patients with heart failure (GSE121893)^14^. After quality filtering for detected genes, we analyzed 9994 cells in normal heart tissues and 4221 cells in heart failure tissues. Four major cell types were identified, including cardiomyocytes, fibroblasts, vascular cells, and immune cells (Fig. S2A-S2D). Cells in failed hearts showed a higher *ACE2*-positive rate than in normal hearts (7.60% of all cells vs. 5.88% of all cells). We observed higher enrichment of *ACE2* expression in cardiomyocytes of failed hearts than that of normal hearts (9.87% of cardiomyocytes vs. 6.75% of cardiomyocytes). In contrast, lower expression of *ACE2* in arterial vascular cells of failed hearts compared to that of the normal (7.93% of arterial vascular cells vs. 19.4% of arterial vascular cells) (Fig. 3A-3B). Immunofluorescence staining revealed a significantly higher percentage of *ACE2*-positive cells in human heart sections of heart failure compared with healthy donors (Fig. 3C).

**Figure 3.**
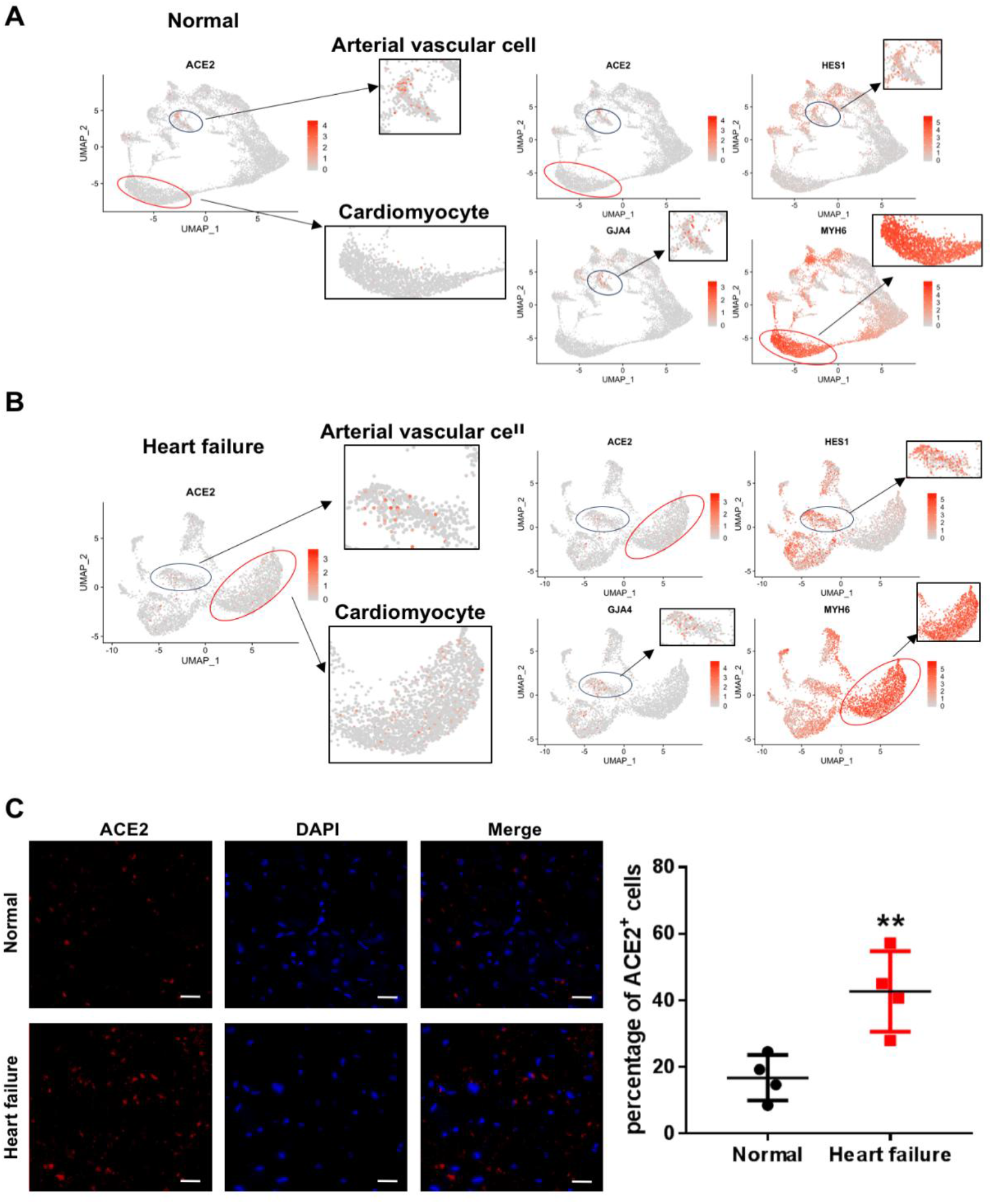
Different distribution of ACE2-expressing cells between normal and failed hearts. A, Feature plot showing the distribution of *ACE2, HES1, MYH6* and *GJA4* expression levels in normal hearts. B, Feature plot showing the distribution of *ACE2, HES1, MYH6* and *GJA4* expression levels in failed hearts. C, Immunofluorescence staining of ACE2 (green) expression and DAPI (blue) in normal hearts, and in failed hearts. Scale bars, 20μm (n=4). Significance is determined with a two-tailed unpaired Student’s t-test, **P<0.01.

Given that *ACE2* gene is highly expressed in arterial vascular cells, we next employed gene ontology (GO) enrichment analysis among *ACE2*-expressing arterial vascular cells (cluster 7, *GJA4*- and *HES1*-positive cells) in the normal hearts. The highly expressed genes in these vascular cells were associated with virus infection response, including viral process, response to the virus, and response to interferon-gamma (Fig. S3A). Kyoto encyclopedia of genes and genomes (KEGG) pathway analysis revealed that these highly expressed genes might be involved in viral myocarditis and pathogenic Escherichia coli infection, indicating that these arterial vascular cells may be the afferent pathway of infectious heart diseases (Fig. S3B). Moreover, these cells highly expressed chemokine (*CXCL12, CCL2)* and integrin subunit (*ITGA1, ITGA7*) (Fig. S3C), which may participate in viral infection, post-infection immune response, migration, and adhesion of immune cells. GO and KEGG analysis of *ACE2*-expressing arterial vascular cells in failed heart also suggested that these cells were highly correlated with virus response and infectious diseases (Fig. S4A-S4B). Unlike normal heart, these cells in failed heart expressed some regulators of monocyte and B cell differentiation, such as *SOX4* and *MEF2C* (Fig. S4C), which might be related to immune response and inflammation.

### Higher gene expression of ACE2 in cells infected by RSV or MERS-CoV

Given that hospital-related transmission of SARS-CoV-2 was suspected in 41% of patients^12^, we speculated that patients simultaneously infected by other viruses may be more susceptible to SARS-CoV-2 infection. We then determined whether other viral infection might affect the expression of ACE2. We analyzed ACE2 expression in cells infected by various viruses, using published RNA-Seq data (GSE140226 and GSE139516)^15^. As expected, *ACE2* expression was significantly up-regulated when lung carcinoma cells were infected with the respiratory syncytial virus and MERS-CoV (Fig. 4A-4B). Moreover, *Ace2* expression in pulmonary endothelial cells of mice was up-regulated 72 hours after lipopolysaccharide (LPS) injection^16^ (Fig. 4C). Together, these results indicate that patients with a virus infection or inflammation might be more susceptible to SARS-CoV-2 infection.

**Figure 4.**
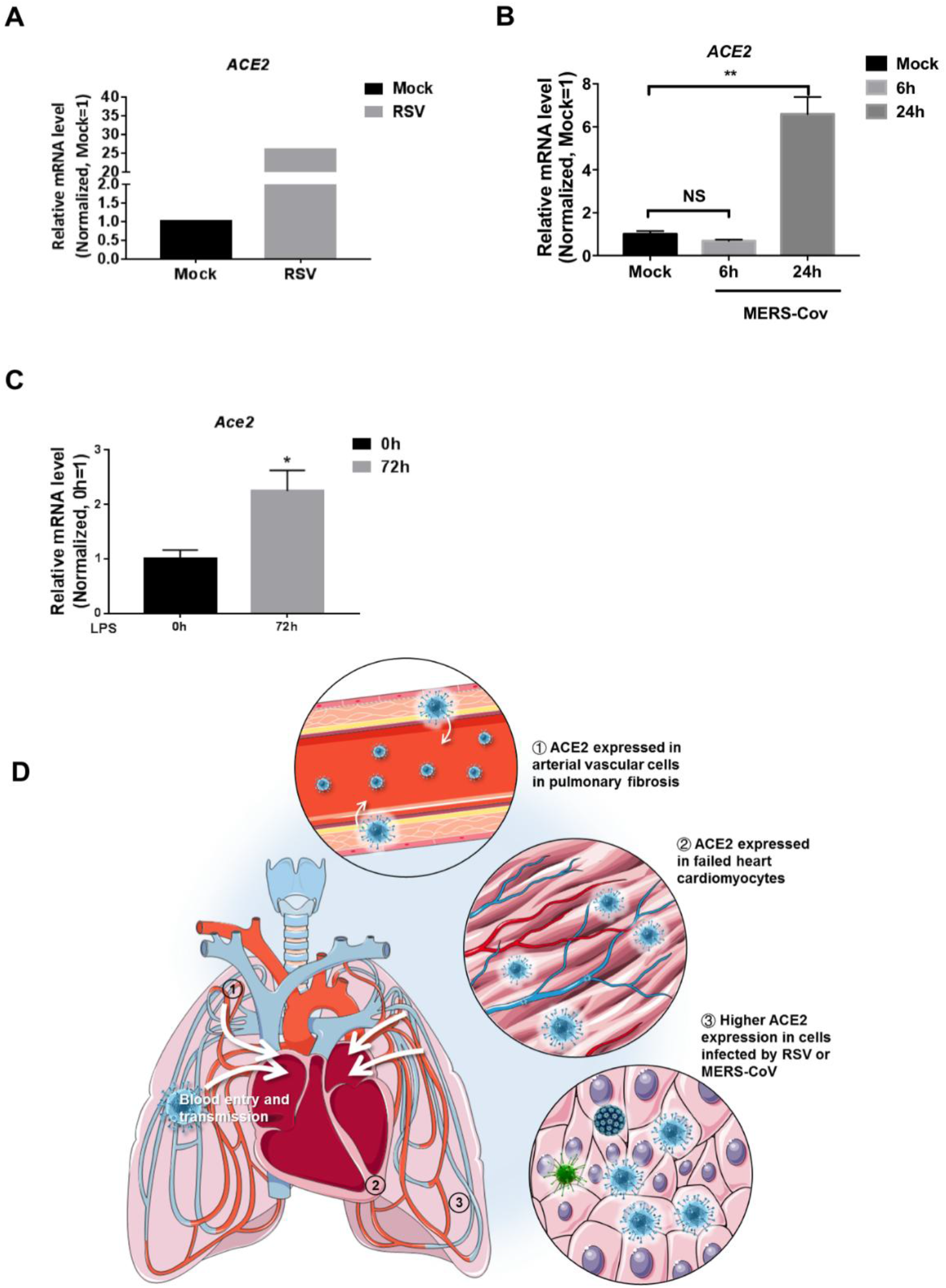
Expression of ACE2 under other infections. A, Expression of *ACE2* in H929 cells treated with mock virus or respiratory syncytial virus (RSV). B, Expression of *ACE2* in Calu-3 cells treated with mock or MERS-CoV 6hpi and 24hpi. (n=3). Significance is determined with one-way ANOVA test, **P<0.01. C, Expression of *Ace2* in lung endothelial cells 0h and 72h after LPS injection in mouse (n=10 in 0h and n=12 in 72h). Significance is determined with two-tailed unpaired Student’s t-test, *P<0.05. D, Schematic review of ACE2 expression providing insight into SARS-CoV-2 blood entry and heart injury.

## Discussion

SARS-CoV-2 has been reported to exist in human feces, urine and blood^17^. Given that very few viruses can survive from gastric acid, the SARS-CoV-2 found in feces is less likely to be transmitted through the digestive tract then through the bloodstream. Urine is also directly filtered from blood. Therefore, the blood entry of SARS-CoV-2 is a pivotal step for its spread to other organs, body fluids and excreta, but how this virus enters bloodstream has not been fully elucidated. ACE2 was shown to be a receptor for SARS-CoV-2, whose expression pattern might provide possible routes for SARS-CoV-2 entry. In the present study, we demonstrated that ACE2 was mainly expressed in arterial vascular cells in fibrotic lungs, which might cause blood transmission of SARS-CoV-2. Patients with COPD bear a high mortality rate after being infected with SARS-CoV-2^12^, and the lungs of COVID-19 patient displayed characteristics of pulmonary fibrosis^18 8^. Therefore, we speculate that pulmonary artery vascular cells may be the main target of SARS-CoV-2 attack in patients with pulmonary fibrosis. SARS-CoV-2 might invade artery vascular cells, recruit immune cells, leading to subsequent inflammatory storms, then enter the bloodstream through the pulmonary artery and cause multiple organs injury in critical patients or patients with underlying lung diseases. Further and more detailed work will be needed to explore whether and how SARS-CoV-2 invades pulmonary artery vascular cells in COVID-19 patients. We also found that *FURIN* but not *TMPRSS2* was highly expressed in pulmonary artery vascular cells, indicating that blocking FURIN may help to prevent SARS-CoV-2 invading the pulmonary artery. Thus, our study has provided new insights into SARS-CoV-2 transmission between organs in critical patients or patients with underlying lung diseases.

The first destination of pulmonary circulation outflow is the heart. Therefore, we assume that SARS-CoV-2 may attack the heart through the blood flow, which may explain the high incidence of cardiac injury in critical patients^12^. Also, heart failure samples displayed a more substantial proportion of ACE2 positive cells in cardiomyocytes and higher overall expression of ACE2 than the normal heart, which might explain why patients with underlying heart diseases were correlated with higher mortality. Thus, preventing SARS-CoV-2 from entering the bloodstream may reduce the chance of myocardial damage in patients with underlying heart diseases. In contrast, the lower proportion of ACE2-expressing cells in arterial vascular cells of failed hearts might be correlated with weaker anti-infection ability, along with the activation of the inflammatory response induced by SARS-CoV-2 infection.

Finally, we observed a higher gene expression of *ACE2* in cells infected by RSV or MERS-CoV. We speculated that patients simultaneously infected by other viruses may have higher expression of *ACE2*, indicating those who bear a coinfection are possibly at a higher risk and are more susceptible to SARS-CoV-2 infection. It may explain the high incidence of hospital-related transmission/infection of SARS-CoV-2^12^. In conclusion, our findings indicate that patients with pulmonary fibrosis, heart failure, and virus infection have a higher risk and are more susceptible to SARS-CoV-2 infection. SARS-CoV-2 might spread into the bloodstream to attack other organs (Fig. 4D). This work provides new insights into how the virus transmits between organs. We also propose a potential therapeutic strategy to mitigate SARS-CoV-2 transmission to the heart and to prevent developing into severe cases in COVID-19 patients.

## Methods

### Cell filtering and cell-type clustering analysis

R package Seurat were used for cell filtration, clustering analysis, and uniform manifold approximation and projection (UMAP) as described in the vignettes (https://satijalab.org/seurat/vignettes.html). Cells expressing <200 or >3000 genes in normal lung samples, <200 or >4000 genes in lung fibrosis samples, <200 or >7000 genes in normal heart sample, and <200 or >5000 genes in heart failure sample were filtered out for exclusion of noncell or cell aggregates. Next, data were log-normalized and identified highly variable genes using the FindVariableFeatures function. ScaleData function were used for linear regression and Principle component analysis (PCA) was performed on the scaled data for linear dimensional reduction. Then, a graph-based clustering approach (FindClusters function) was performed to cluster cells and Uniform Manifold Approximation and Projection (UMAP) was used to visualize clusters. Visualization of gene expression with violin plot, feature plot was implemented by VlnPlot, FeaturePlot function. FindAllMarkers function (min.pct = 0.25, logfc. threshold = 0.25) was used to identify marker genes. Type II alveolar cell, endothelial cell, smooth muscle cell, immunocyte, ciliated cell, fibroblast, and cardiomyocyte clusters were removed based on their expression of known type II alveolar cell markers such as *SFTPC*, endothelial cell markers such as *PECAM1, VWF* and *GJA4*, smooth muscle cell markers such as *ACTA2* and *MYH11*, immunocyte markers such as *CD163, HLA-DRA, GNLY, CD68* and *PTPRC*, ciliated cell markers such as FCN3 and TPPP3, fibroblast markers such as *DCN* and *COL1A2*, and cardiomyocyte markers such as *MYH6, MYH7* and *TNNT2*. Gene ontology (GO) and Kyoto encyclopedia of genes, genomes (KEGG) pathway analysis were performed with marker genes on DAVID website (https://david.ncifcrf.gov/).

### Immunofluorescence

The normal heart and heart failure samples were permeabilized with 0.3% Triton X-100 for 10 min and blocked with 5% donkey serum in PBS for 2h, incubated with primary antibodies ACE2 (1:50) (affinity #AF5165) overnight at 4°C, washed with PBS, incubated with secondary antibodies for 1h at room temperature, washed with PBS and then stained with DAPI.

### Clinical specimens

We collected 4 cases of normal heart and 4 cases of heart failure where the death occurred suddenly and unexpectedly. Reviewing patients’ charts were exempted from informed consent by the Ethics Committee Board at the School of Basic Medical Sciences, Fudan University.

### Statistical analysis

Data are presented as mean ± SEM. Statistical analyses were conducted with GraphPad prism 7. Differences among groups were determined with one-way ANOVA followed by Tukey test for multiple comparisons. Differences between two groups were determined with unpaired Student’s t-test.

## Data availability

Public data for normal and fibrotic lungs scRNA-seq is available in GSE132771. Public data for healthy hearts scRNA-seq was available in GSE109816, failed hearts scRNA-seq in GSE121893. Public data for respiratory syncytial virus infection RNA-seq is available in GSE140226, MERS-CoV in GSE139516, and LPS injection in GSE136848.

## Funding

This work was supported by the Shanghai Science and Technology Commission of China (19JC1411300 to D. Meng), General Programs (81873469 to D. Meng, 81873536 to X. Wang, 81572713 to X. Zhi), and the Great Program (91639103 to D. Meng) of the National Natural Science Foundation of China, and China Postdoctoral Science Foundation (2019M651371 and 2019T120303 to X. Wei).

## Author contributions

M.D. conceived and designed the project. G.J. analyzed most experiments. W.X. performed immunofluorescence staining of ACE2. L.L., Y.Z., S.Y., Q.Y., Z.X., W.X., and Z.X. provided reagents, advice, and discussed results. M.D., G.J., and L.Q. wrote the manuscript. All authors discussed results and commented on the manuscript.

## Competing interests

The authors declare that they have no competing interests.

## Notes

### Competing Interest Statement

The authors have declared no competing interest.

